# Three Year Outcomes after Programmatic Transitioning to Dolutegravir in the Context of Severe Civil Unrest in Haiti

**DOI:** 10.1101/2024.08.13.24311952

**Authors:** Bernard Liautaud, Ana Sanchez Chico, Youry Macius, Patrice Joseph, Harrison Reeder, Theo Bolas, Adias Marcelin, Colette Guiteau Moïse, Alexandra Apollon, Angela Dai, Pierre Cremieux, Jean W. Pape, Serena P. Koenig

**Affiliations:** Haitian Group for the Study of Kaposi’s Sarcoma and Opportunistic Infections (GHESKIO), Port-au-Prince, Haiti; Analysis Group, Boston, Massachusetts, USA; Massachusetts General Hospital, Boston, Massachusetts, USA; Harvard Medical School, Boston, MA, USA; Weill Cornell Medicine, New York, New York, USA; Brigham and Women’s Hospital, Boston, Massachusetts, USA

**Keywords:** ART, HIV care continuum, ART resistance, Integrase inhibitors

## Abstract

**Background:** Dolutegravir (DTG) has been widely scaled-up worldwide. Data on long-term outcomes are limited.

**Methods:** We included all persons living with HIV (PLWH) ≥15 years of age who initiated or switched to DTG in Port-au-Prince, Haiti. We described treatment outcomes by pre-switch viral load and assessed predictors of virologic failure using multivariable logistic regression.

**Findings:** A total of 10,354 PLWH initiated or switched to DTG from November 5, 2018 to March 21, 2021, and were included in the analyses. Of these, 2217 (21.4%) were ART-naïve and 8137 (78.6%) switched from an NNTRI-based regimen. Median follow-up time on DTG was 2.8 years (IQR: 2.3, 3.1). Among PLWH with ≥2 tests on DTG, 83.5% of ART-naïve, 93.1% with pre-switch suppression, 64.8% with pre-switch failure, and 90.2% without pre-switch viral load had <1000 copies/mL at latest test. Among treatment-experienced patients, predictors of HIV-1 RNA ≥1000 copies/mL at latest test included younger age (adjusted odds ratio [aOR]: 2.33 for ≤30 vs. ≥50 years), shorter pre-switch time on ART (aOR: 0.91; 95% CI: 0.89, 0.94/year), lower education (aOR: 1.31; 95% CI: 1.10, 1.55), and higher pre-switch viral load: (aOR: 7.00; 95% CI; 5.74, 8.54 for ≥10,000 vs. <1000 copies/mL).

**Interpretation:** Virologic outcomes on DTG-based regimens are outstanding for PLWH with pre-switch suppression, even in the context of severe civil unrest in Haiti. However, rates of persistent viremia are high among PLWH who experienced pre-switch failure; additional interventions are critically needed, including access to genotypic resistance testing and provision of alternative regimens as clinically-indicated.

**Funding:** None

## INTRODUCTION

Dolutegravir (DTG)-based antiretroviral therapy (ART) regimens are prescribed for over 90% of persons living with HIV (PLWH) worldwide, in accordance with World Health Organization guidelines.^1,2^ DTG offers many advantages over previous ART medications – high potency, a high genetic barrier to resistance, favorable tolerability, and few drug interactions.^3^ It is anticipated that DTG will serve as the primary backbone medication for antiretroviral therapy (ART) regimens in US President’s Emergency Plan for AIDS Relief (PEPFAR) programs for the foreseeable future.^2^ Therefore, it is essential to maximize viral suppression among PLWH, particularly in countries with limited access to drug resistance testing.

As DTG became the preferred ART regimen over the last four years, PLWH in many settings were switched from first-line non-nucleoside reverse transcriptase inhibitor (NNRTI)-based regimens to DTG-based regimens, without regard to viral suppression, and without drug resistance testing for those with virologic failure.^1^ Studies from programmatic settings in low and middle income countries (LMICs) have reported high rates of viral suppression on DTG-based regimens, but data are limited beyond the first-year of treatment.^4–7^ Long-term data on DTG outcomes are also limited from the Latin America and Caribbean region.^4,8^

Pre-switch viremia is associated with future virologic failure on DTG-based regimens, which is attributed to continued poor adherence and/or the presence of nucleoside reverse transcriptase inhibitor (NRTI) mutations.^5,6,9,10^ Recent studies from various African countries have documented the emergence of DTG resistance, particularly in PLWH with virologic failure prior to switching to DTG-based regimens.^11–15^ However, access to resistance testing remains very limited in many LMICs. Furthermore, in settings of severe civil unrest, treatment outcomes may be further compromised by structural barriers to adherence.

In contrast, rates of virologic rebound on DTG-based regimens appears to be low among patients with pre-switch suppression.^5–7,16,17^ Longer-term data on virologic outcomes could provide evidence to support the implementation of DTG-3TC in this low-risk cohort. Studies from higher-income settings have demonstrated that DTG-3TC is non-inferior to standard three-drug therapy in eligible patients, and this strategy is now recommended in US and European guidelines, and is being implemented in some countries in the Latin American and Caribbean region.^18–24^

We conducted a retrospective analysis of longitudinal virologic outcomes for patients who initiated or switched to DTG from a first-line efavirenz (EFV)-based regimen at the Haitian Group for the Study of Kaposi’s Sarcoma and Opportunistic Infections (GHESKIO). We stratified results based on ART history and virologic status at TLD initiation.

## METHODS

### Study Setting and Patient Characteristics

GHESKIO is a Haitian non-governmental organization, and the largest provider of HIV care in the Caribbean. HIV care in Haiti is largely funded by the US President’s Emergency Plan for AIDS Relief PEPFAR and the Global Fund to Fight AIDS, Tuberculosis, and Malaria, and all care at GHESKIO is provided free of charge. The estimated adult HIV prevalence in Haiti is 1.7%.^25^ Haiti is ranked 163 of 191 countries on the Human Development Index, and during the study period, Haiti was experiencing a period of severe political instability and gang-related violence.^26^ Over 80% of the capital city of Port-au-Prince is controlled by armed gangs, and GHESKIO’s central facilities are located adjacent to some of the country’s largest and most impoverished slums, where gang violence is most intense.^27^

GHESKIO has been providing ART since 2003, and until 2017, viral load monitoring was not routinely available. First-line therapy included an NNRTI-based regimen in combination with two NRTI’s; in 2010 the preferred regimen was EFV/tenofovir disoproxil fumarate (TDF)/3TC. In 2018, the TDF/3TC/DTG (TLD) regimen became available, and most patients taking other regimens switched to TLD, regardless of viral suppression.

### Study Design

We conducted a retrospective cohort analysis of all PLWH ≥15 years of age at ART initiation who initiated or switched to TLD at GHESKIO from November 5, 2018 to March 21, 2021, had at least one HIV-1 RNA test after starting TLD, and were ART-naïve at TLD initiation or switched to TLD from a first-line NNRTI-based regimen. Patients were followed until March 21, 2022, the date of database closure.

Patients’ baseline status was classified by ART regimen and pre-switch HIV-1 RNA <1000 copies/mL or ≥1000 copies/mL at TLD initiation; those with no HIV-1 RNA measurement within 180 days prior to switching were classified as having no available viral load at TLD switch. A threshold of 1000 copies/mL was used to define virologic failure, in accordance with WHO guidelines, because dried blood spot testing is used for GHESKIO patients who receive viral load testing at community HIV clinics.^28^

The outcome of patient retention was defined according to status at database closure. Patients were considered active if they were no more than 90 days late for their most recent appointment at the time of database closure; they were considered late if they were 91 to 365 days late for their most recent appointment, and as lost to follow-up (LTFU) if they were >365 days late. Patients were classified as dead if they were known to have died and transferred if they were known to have transferred to a different ART provider.

The outcome of patient viral load at follow-up testing was also dichotomized as HIV-1 RNA ≥1000 versus <1000, the same threshold as at study baseline. The main outcome of interest was dichotomized viral load at the most recent test. Longitudinal dichotomized viral load measurements for those with multiple tests after TLD initiation were also examined as described below. It was not feasible to include confirmatory viral load results, because there was variability in the timing and completion rate of confirmatory viral load testing.

### Statistical Analysis

Data on sex, age, education, marital status, ART regimen at time of TLD initiation, time on ART at TLD initiation, and all HIV-1 RNA results were extracted from GHESKIO’s electronic medical record (EMR). Baseline characteristics were summarized in the overall cohort using frequencies and proportions for categorical variables and medians with interquartile ranges (IQR) for continuous variables.

We summarized patient retention outcomes by ART regimen and viral load status at start of TLD. Time from TLD initiation to patient loss (either LTFU or death) was assessed using Kaplan-Meier curves, treating patient transfer and database closure as censoring mechanisms.

We also described the proportion of patients with HIV-1 RNA ≥1000 at most recent test, stratified by ART regimen and virologic status at TLD initiation. Among active patients who had received at least two viral load tests, we calculated the proportion with virologic failure on the first and latest HIV-1 RNA test.

Finally, we identified predictors of having HIV-1 RNA ≥1000 copies/mL at most recent test among active patients, using a multivariable logistic regression model based on observed characteristics of the patients at TLD initiation, namely age, sex, education, civil status, residence location, time on ART, and virologic status. Data were analyzed using Stata version 16 software (StataCorp LLC, College Station, TX).

### Ethical Considerations

Due to the retrospective study design, it was not feasible to obtain informed consent. This study was reviewed and approved by the institutional review boards of GHESKIO, Weill Cornell Medical College, and Brigham and Women’s Hospital.

## RESULTS

A total of 10,932 PLWH ≥15 years of age initiated TLD or switched to TLD from an NNRTI-based first-line regimen at GHESKIO from November 5, 2018 to March 21, 2021. Of these, 10,354 (94.7%) had at least one viral load test after starting TLD and were included in the analyses (see **Table 1**). The median age at TLD initiation was 42.7 years (IQR: 34.6, 51.4), 6054 (58.5%) were female, and 5053 (48.8%) had no education or primary school only (see **Table 1**).

**Table 1.** Baseline Characteristics (n=10,354)

| Variable |  |
| --- | --- |
| Female Sex – no. (%) | 6054 (58.5%) |
| Age at TLD initiation – median (IQR) | 42.7 (34.6, 51.4) |
| Education – no. (%) |  |
| None | 1809 (17.5%) |
| Primary school | 3244 (31.3%) |
| Secondary or higher | 4907 (47.4%) |
| Missing data | 394 (3.8%) |
| Marital Status |  |
| Married or living together | 5079 (49.1%) |
| Formerly married | 1832 (17.7%) |
| Single | 3060 (29.6%) |
| Missing data | 383 (3.7%) |
| Residence Zone in Port-au-Prince |  |
| Carrefour | 2303 (22.2%) |
| Delmas | 1427 (13.8%) |
| Plaine | 2097 (20.3%) |
| Downtown Port-au-Prince | 2314 (22.3%) |
| Petionville | 799 (7.7%) |
| Outside of metropolitan Port-au-Prince | 1414 (13.7%) |
| ART Regimen and Virologic Status at Time of Start or Switch to TLD – no. (%) |  |
| ART-naïve | 2217 (21.4%) |
| Pre-switch HIV-1 RNA <1000 copies/mL on first-line NNRTI-based regimen | 5195 (50.2%) |
| Pre-switch HIV-1 RNA ≥1000 copies/mL on first-line NNRTI-based regimen | 1493 (14.4%) |
| First-line NNRTI-based regimen without recent HIV-1 RNA test | 1449 (14%) |
| Time on ART Regimen at Switch to TLD– years (IQR) |  |
| Pre-switch HIV-1 RNA <1000 copies/mL on first-line NNRTI-based regimen | 4.6 (2.4, 8.2) |
| Pre-switch HIV-1 RNA ≥1000 copies/mL on first-line NNRTI-based regimen | 3.9 (2.3, 6.1) |
| First-line NNRTI-based regimen without recent HIV-1 RNA test | 4.3 (2.4, 7.4) |

A total of 2217 (21.4%) were ART-naïve and 8137 (78.6%) switched from a first-line NNTRI-based regimen. Among the patients switching to TLD, 1493 (18.3%) had pre-switch HIV-1 RNA ≥1000 copies/mL, 5195 (63.8%) had pre-switch HIV-1 RNA <1000 copies/mL, and 1449 (17.8%) had no pre-switch viral load measurement. The median time on ART was 4.37 years (IQR: 2.37, 7.49) at time of switch to TLD (see **Table 1**).

Among the total cohort, the median observed follow-up time on TLD was 2.8 years (IQR: 2.3, 3.1). At time of study database closure, 8728 (84.3%) were classified as active, 440 (4.2%) had transferred, 511 (4.9%) were late for their final visit, 576 (5.6%) were LTFU, and 99 (1.1%) had died (see **Figure 1**). A lower proportion of patients with pre-switch HIV-1 RNA ≥1000 copies/mL had active status at the end of the study period. **Figure 2** is a Kaplan-Meier Curve of retention in care by switch category, with the exclusion of patients who were transferred.

**Figure 1.**
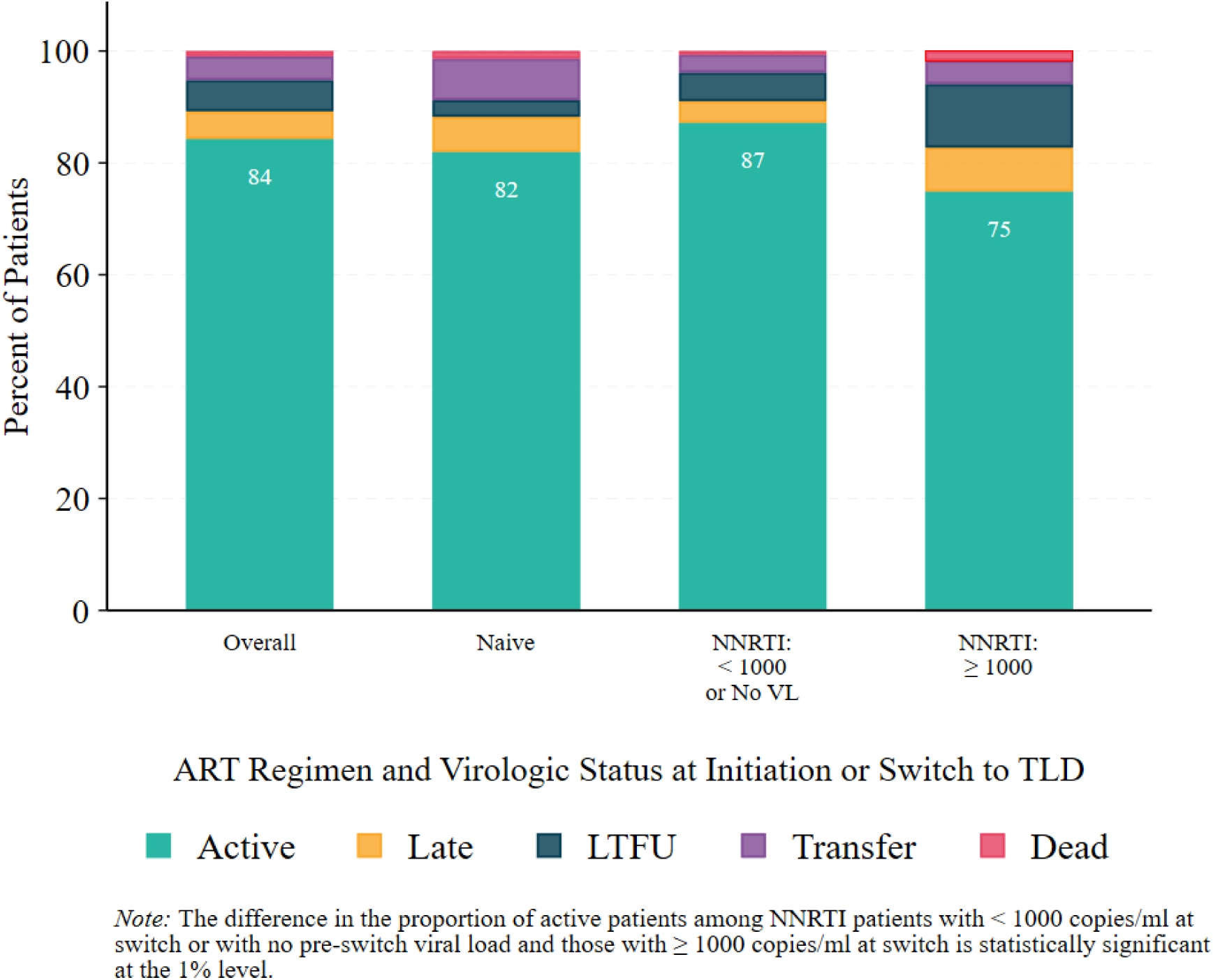
Final Outcome by ART Regimen and Virologic Status at Start of TLD.

**Figure 2.**
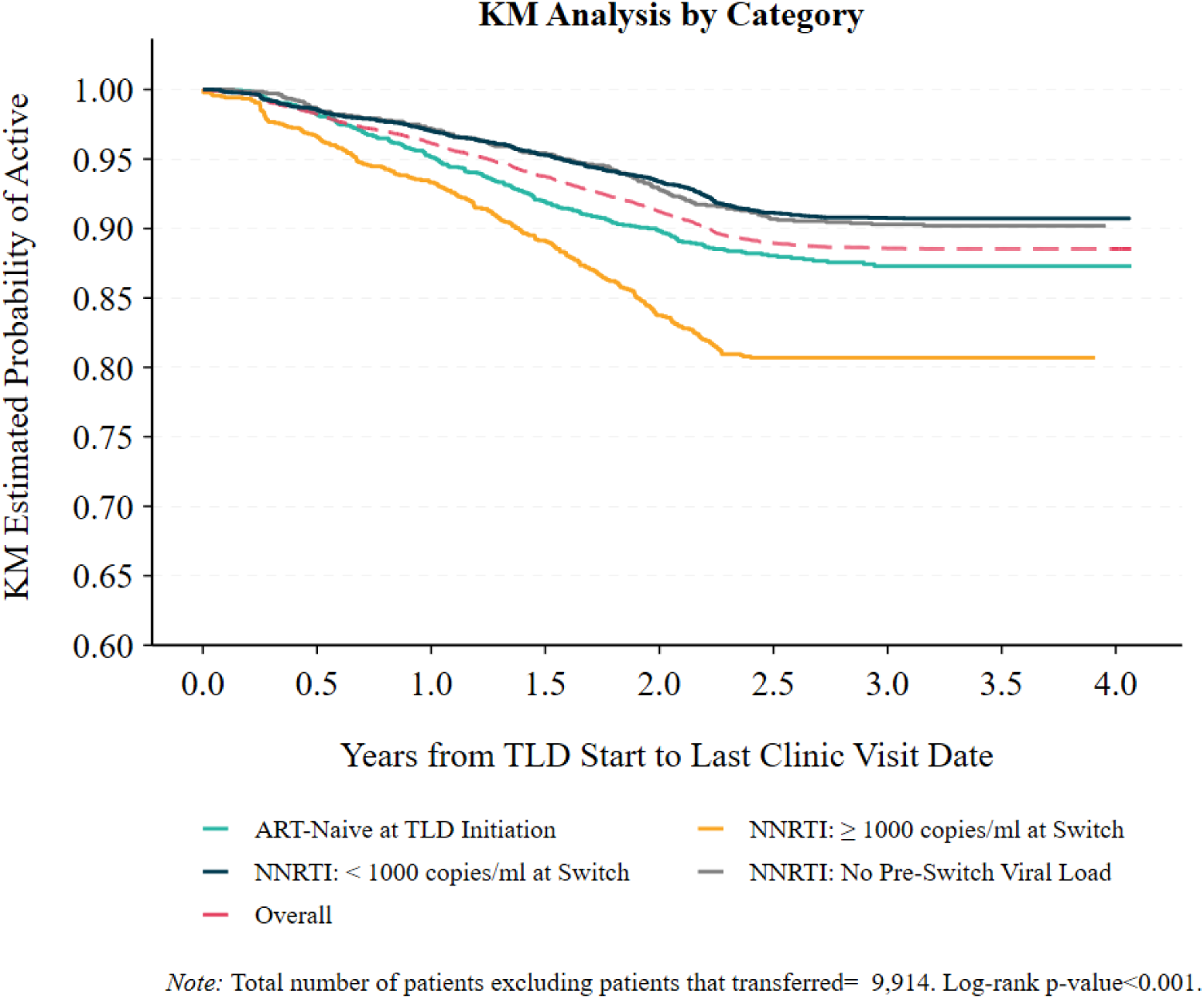
Kaplan Meier Curve, by Virologic Status at Start of TLD.

Among the 8728 active patients, 7597 (87.0%) had HIV-1 RNA <1000 copies/mL at their latest test before database closure; these proportions were 83.5% (1518/1817) among those who were ART-naïve at TLD initiation, 93.1% (4215/4529) in those with pre-switch HIV-1 RNA <1000 copies/mL, 64.8% (724/1118) in those with pre-switch HIV-1 RNA ≥1000 copies/mL and 90.2% (1140/1264) in those without a pre-switch viral load (see **Table 2**). The proportion of patients with HIV-1 RNA <1000 copies/mL at latest test was lower in inactive patients (late, LTFU, transferred, or died), including 73.7% of those who were ART-naïve at TLD initiation and 72.8% among those switching from NNRTI-based regimens.

**Table 2.**
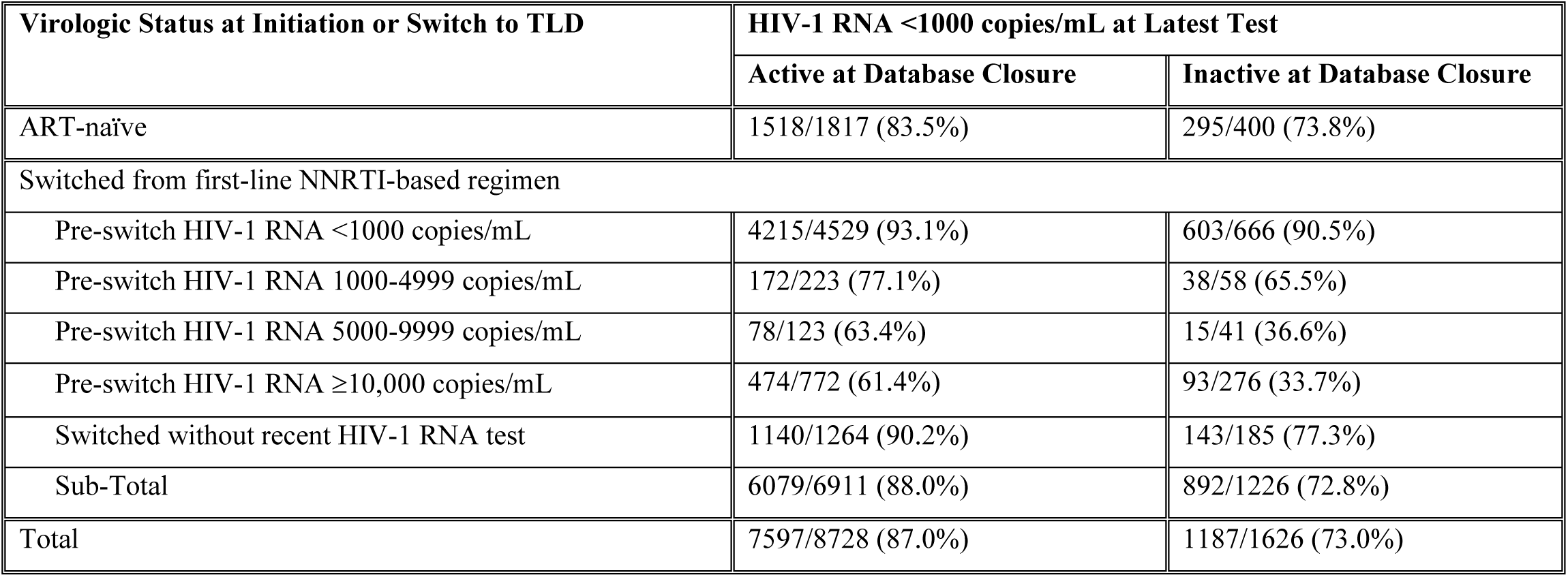
Proportion of Patients on TLD with HIV-1 RNA <1000 copies/mL at Latest Test.

| Virologic Status at Initiation or Switch to TLD | HIV-1 RNA <1000 copies/mL at Latest Test |  |
| --- | --- | --- |
|  | Active at Database Closure | Inactive at Database Closure |
| ART-naïve | 1518/1817 (83.5%) | 295/400 (73.8%) |
| Switched from first-line NNRTI-based regimen |  |  |
| Pre-switch HIV-1 RNA <1000 copies/mL | 4215/4529 (93.1%) | 603/666 (90.5%) |
| Pre-switch HIV-1 RNA 1000-4999 copies/mL | 172/223 (77.1%) | 38/58 (65.5%) |
| Pre-switch HIV-1 RNA 5000-9999 copies/mL | 78/123 (63.4%) | 15/41 (36.6%) |
| Pre-switch HIV-1 RNA ≥10,000 copies/mL | 474/772 (61.4%) | 93/276 (33.7%) |
| Switched without recent HIV-1 RNA test | 1140/1264 (90.2%) | 143/185 (77.3%) |
| Sub-Total | 6079/6911 (88.0%) | 892/1226 (72.8%) |
| Total | 7597/8728 (87.0%) | 1187/1626 (73.0%) |

Among active patients, 80.8% (1469/1817) of ART-naïve, 93.2% (4222/4529) with pre-switch HIV-1 RNA <1000 copies/mL, 82.8% (926/1118) with pre-switch HIV-1 RNA ≥1000 copies/mL, and 94.0% (1188/1264) of patients without a recent pre-switch viral load had at least two viral load tests after initiating TLD. Among patients with at least two tests on TLD who were ART-naïve at TLD initiation, 82.6% had viral suppression on their latest test. A total of 74.1% had viral suppression on both their first and latest test on TLD; 18.9% had viral suppression on one of the two tests, and 7.0% had failure on both tests (see **Figure 3**).

**Figure 3.**
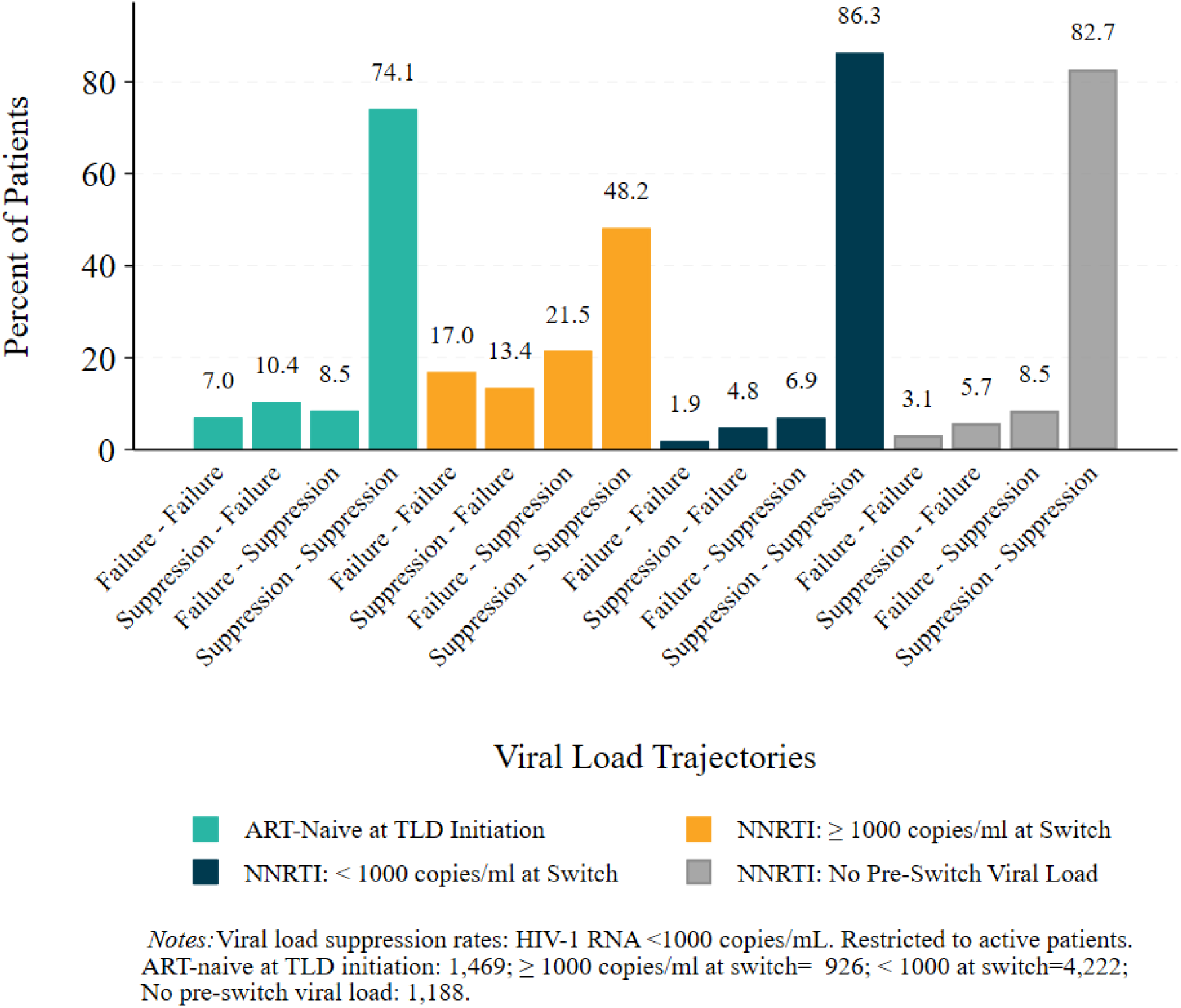
First and Latest Viral Load Results after Switch to TLD, among Active Patients*.

Among active patients with pre-switch HIV-1 RNA <1000 copies/mL and at least two tests on TLD, 93.2% had viral suppression on their latest test (see **Figure 3**). A total of 86.3% had viral suppression on both their first and latest test; 11.7% had viral suppression on one of the two tests, and 1.9% had failure on both their first and latest test on TLD. Among the 3847 patients with pre-switch HIV-1 RNA <1000 copies/mL who also had virologic suppression at first test on TLD, 3644 (94.7%) were still suppressed at most recent viral load test.

Among patients with pre-switch HIV-1 RNA ≥1000 copies/mL and at least two tests on TLD, 69.7% had viral suppression on their latest test. A total of 48.2% had viral suppression on both their first and latest test; 34.9% had viral suppression on one of the two tests, and 17.0% had failure on both their first and latest test on TLD (see **Figure 3**). Among patients without a recent pre-switch viral load, 91.2% had viral suppression on their latest test. A total of 82.7% had viral suppression on both their first and latest test; 14.2% had viral suppression on one of the two tests, and 3.1% had failure on both their first and latest test on TLD.

In multivariable analyses limited to active patients, predictors of virologic failure at latest test included higher pre-switch viral load or no recent pre-switch viral load, compared with pre-switch HIV-1 RNA <1000 copies/mL (≥10,000 copies/mL: adjusted odds ratio (aOR) 7.00 [95% CI: 5.74, 8.54]; 5000 to 9999 copies/mL: aOR 6.21 [95% CI: 4.05, 9.50]; 1000 to 4999 copies/mL: aOR 3.08 [95% CI: 2.15, 4.41]; no recent pre-switch viral load: aOR 1.49 [95% CI: 1.19, 1.86]) and no education or primary school only (OR: 1.31; 95% CI: 1.10, 1.55). Older age (30 to 50 vs. <30 years: aOR 0.68 [95% CI: 0.53, 0.87]; ≥50 vs. <30 years: aOR 0.43 [95% CI: 0.32, 0.58]), and longer time on ART at switch to TLD (aOR: 0.91/year [(95% CI: 0.89, 0.94]) were associated with a lower risk of virologic failure (see **Table 3**).

**Table 3.** Univariable and Multivariable Predictors of HIV-1 RNA ≥1000 Copies/mL at Latest Test, Among Active Patients who Switched to TLD.

| Variable | Univariable Analysis |  | Multivariable Analysis |  |
| --- | --- | --- | --- | --- |
|  | Odds Ratio (95% CI) | p-value | Adjusted Odds Ratio (95% CI) | p-value |
| Male sex (vs. female) | 1.05 | 0.542 | 1.12 (0.95 - 1.33) | 0.177 |
| Age 30 to 50 years (vs. ≤30) | 0.55 | <0.001 | 0.68 (0.53 - 0.87) | 0.002 |
| Age ≥50 years (vs. ≤30) | 0.28 | <0.001 | 0.43 (0.32 - 0.58) | <0.001 |
| No education or primary only (vs. at least some secondary) | 1.22 | 0.011 | 1.31 (1.10 - 1.55) | 0.002 |
| Married (vs. single/formerly married) | 0.90 | 0.182 | 1.02 (0.86 - 1.20) | 0.836 |
| Residence Zone (vs. Carrefour Zone) |  |  |  |  |
| Downtown Port-au-Prince | 1.05 | 0.672 | 0.93 (0.73 - 1.19) | 0.580 |
| Delmas | 0.80 | 0.091 | 0.89 (0.67 - 1.17) | 0.388 |
| Plaine | 0.85 | 0.156 | 0.83 (0.65 - 1.07) | 0.149 |
| Petionville | 0.74 | 0.078 | 0.72 (0.50 - 1.03) | 0.072 |
| Outside of Port-au-Prince | 1.16 | 0.227 | 1.31 (1.01 - 1.70) | 0.044 |
| Time on ART at Switch to TLD (years) | 0.89 | <0.001 | 0.91 (0.89 - 0.94) | <0.001 |
| Pre-switch HIV-1 RNA (vs. <1000 copies/mL) |  |  |  |  |
| No recent HIV-1 RNA at switch | 1.44 | 0.001 | 1.49 (1.19 - 1.86) | <0.001 |
| 1000 to 4999 copies/mL | 3.38 | <0.001 | 3.08 (2.15 - 4.41) | <0.001 |
| 5000 to 9999 copies/mL | 6.44 | <0.001s | 6.21 (4.05 - 9.50) | <0.001 |
| ≥10,000 copies/mL | 7.31 | <0.001 | 7.00 (5.74 - 8.54) | <0.001 |

## DISCUSSION

This study assessed 3-year virologic outcomes after the switch to TLD in the context of severe civil unrest in Haiti. Though the socio-political situation has presented extraordinary challenges to the health system, the overwhelming majority of patients who were suppressed at switch to TLD maintained viral suppression. In contrast, patients with pre-switch viremia were more likely to drop out of care, and among those who remained active, about one-third had HIV-1 RNA ≥1000 copies/mL on their most recent test. Furthermore, the magnitude of viremia at switch to TLD was associated with future virologic failure on TLD. This high-risk subgroup of patients with detectable pre-switch viremia and persistent viremia on TLD should be targeted for more intensive adherence interventions and access to DTG resistance testing.

Though data from other sites in the Latin America and Caribbean region are limited, studies from various African countries have also reported an association between pre-switch viremia and future virologic failure on TLD. A study from five African countries in the International epidemiology Databases to Evaluate AIDS (IeDEA) research consortium found that patients who switched to DTG with HIV-1 RNA ≥1000 copies/mL had worse HIV treatment outcomes at median follow-up time of 11 months, compared with those who had pre-switch suppression.^6^ Cohort studies from Malawi and Lesotho reported that though viremia after switching to TLD was rare, it was associated with pre-switch viremia.^5,10^ Furthermore, as we found in our study, the risk of viremia increased with higher pre-switch viral loads. A cohort study from South Africa reported rates of viral suppression of about 60% in patients who switched to DTG from a failing NNRTI-based regimen, with lower rates of suppression in those with pre-switch HIV-1 ≥10,000 copies/mL.^9^ These findings emphasize the importance of assessing the magnitude of pre-switch viremia, as well as reporting binary outcomes with discrete viral load thresholds.

We also found that younger age, lower education, and a shorter time on ART prior to switching treatment were associated with virologic failure on TLD; these characteristics have also been associated with poor outcomes in other GHESKIO studies.^29,30^ Younger age was also associated with poor outcomes in a study of second-line DTG-based ART in South Africa, and with viremia and disengagement from care among patients on DTG in the IeDEA consortium.^6,9^ It is likely that patients with pre-switch viremia are a high-risk subgroup, who are more likely to experience challenges to adherence before and after switch to TLD. Demographic and social determinants are also associated with adherence among ART-naïve patients who initiate DTG-based regimens. In the ADVANCE trial, younger age and unemployment were predictors of lower ART adherence, and adherence was correlated with viral suppression.^31^

The majority of patients with persistent viremia on TLD have susceptible virus, demonstrating that adherence interventions are essential.^5,17,32,33^ However, emerging data demonstrate that INSTI-associated drug resistance mutations are detected in a subgroup of patients with virologic failure on DTG-based regimens.^17,34^ A recently published rapid scoping review concluded that among ART-experienced PLWH receiving a second-line DTG-based regimen, genotypic resistance testing may be beneficial to inform treatment changes.^11^ In Lesotho, rates of virologic failure on DTG were low, but DTG resistance mutations were detected in nearly 10% of patients with pre-switch viremia.^10^ Four laboratory-based studies from Africa reported rates of integrase strand transfer inhibitor (INSTI) mutations ranging from 3 to 21%.^12–15^ However, data on emergent DTG resistance in programmatic settings in the Latin America and Caribbean region, with a predominance of subtype B virus, are limited. To date, no systemic surveillance testing for DTG resistance has been published among adults in Haiti, but a case of DTG resistance was detected in an ART-naïve infant identified during a national drug resistance survey conducted through the country’s early infant diagnosis program.^35^

Our study also provides longer-term follow-up data supporting the assertion that patients with pre-switch suppression continue to maintain viral suppression after switch to TLD. The DO-REAL study from Lesotho reported 4-month viral suppression rates of 98%.^36^ A cohort of patients from Malawi reported that viral suppression was maintained at 12 months after switching to TLD.^5^ In the AFRICOS cohort study, 94% of patients remained suppressed at their first viral load on TLD.^16^ Furthermore, long-term data from the ADVANCE trial demonstrated that patients with virologic failure on DTG-based regimens were often able to re-suppress without changing regimen.^32^

Clinical trials conducted in ART-naïve and treatment-experienced patients at low risk of DTG or 3TC resistance have demonstrated that dual therapy is non-inferior to standard three-drug regimens.^20–23^ Several countries in Latin America, including Argentina and Brazil, are routinely implementing DTG-3TC.^24^ However, DTG-3TC is not available in PEPFAR programs. Switching from TLD to TD-3TC would avert TDF-associated renal and bone toxicity.^37,38^ This would require testing for hepatitis B, but inexpensive, point-of-care tests for hepatitis B surface antigen are now available.^39^ The benefits of a TDF-sparing regimen may be even greater in LMICs, with limited capacity to monitor for renal function, lack of access to alternative regimens, and minimal options for those who develop renal failure.

In order to achieve the 95-95-95 targets of the Joint United Nations Programme on HIV/AIDS (UNAIDS) in the Latin America and Caribbean region, it will be necessary to provide HIV services in settings of civil and political unrest.^40^ Our findings demonstrate that GHESKIO’s HIV program remains highly functional, in spite of extraordinary challenges. With nearly three years of follow-up, about 85% of PLWH were active in care, and of these, 87% had HIV-1 RNA <1000 copies/mL at most recent testing. Data on outcomes from other settings of conflict in the region are limited, but in the Central African Republic and Yemen, multi-month medication dispensing and community-based ART sites have been found to be effective for the provision of HIV care.^41,42^ GHESKIO also implements these strategies to ensure ongoing access to ART.

Our study is limited by being conducted in a large urban clinic, which may limit generalizability to other settings. In addition, we were not able to categorize tests with virologic failure as initial or confirmatory testing. Furthermore, this analysis does not include adherence measures.

## CONCLUSIONS

In conclusion, virologic outcomes on TLD are outstanding for PLWH with pre-switch suppression, even in the context of severe civil unrest in Haiti. However, rates of persistent viremia are high among PLWH who experienced pre-switch failure. Additional interventions are critically needed for PLWH and persistent viremia on TLD, including access to resistance testing and alternative treatment options for those in whom DTG resistance is detected.

## Data Availability

Anonymized participant data used in this data will be made available upon request to the corresponding author.

## NOTES

### Contributors

Conceptualization and Methodology: BL, AS, PJ, PC, JWP, SPK; Formal Analysis, AS, HR, AD, PC; Data curation: BL, AS, YM, PJ, HR, AM, AA, AD; Writing (original draft): AS, SPK. Writing (review and editing): BL, YM, PJ, HR, TB, AM, CGM, AA, AD, PC, JWP. Graphics. Supervision: AS, HR. All authors were granted access to the study data and contributed to the decision to submit the study for publication.

## Acknowledgements

PEPFAR and the Global Fund to Fight AIDS, Tuberculosis and Malaria provide funding for HIV service delivery at GHESKIO, but they had no role in the study design, data collection, data analysis, interpretation, or writing of this report.

## REFERENCES

1. Consolidated Guidelines on HIV Prevention, Testing, Treatment, Service Delivery and Monitoring: Recommendations for a Public Health Approach, July 2021.

2. PEPFAR 2023 Country and Regional Operational Plan (COP/ROP) Guidance for all PEPFAR-Supported Countries. Accessed April 12, 2024 at: https://www.state.gov/wp-content/uploads/2023/02/PEPFAR-2023-Country-and-Regional-Operational-Plan.pdf.

3. Venter WDF, Moorhouse M, Sokhela S, et al. Dolutegravir plus Two Different Prodrugs of Tenofovir to Treat HIV. N Engl J Med. Aug 29 2019;381(9):803–815. doi:10.1056/NEJMoa1902824

4. Meireles MV, Pascom ARP, Duarte EC, McFarland W. Comparative effectiveness of first-line antiretroviral therapy: results from a large real-world cohort after the implementation of dolutegravir. AIDS. Aug 1 2019;33(10):1663–1668. doi:10.1097/QAD.0000000000002254

5. Schramm B, Temfack E, Descamps D, et al. Viral suppression and HIV-1 drug resistance 1 year after pragmatic transitioning to dolutegravir first-line therapy in Malawi: a prospective cohort study. Lancet HIV. Aug 2022;9(8):e544–e553. doi:10.1016/S2352-3018(22)00136-9

6. Romo ML, Edwards JK, Semeere AS, et al. Viral Load Status Before Switching to Dolutegravir-Containing Antiretroviral Therapy and Associations With Human Immunodeficiency Virus Treatment Outcomes in Sub-Saharan Africa. Clin Infect Dis. Sep 10 2022;75(4):630–637. doi:10.1093/cid/ciab1006

7. Dorward J, Sookrajh Y, Khubone T, et al. Implementation and outcomes of dolutegravir-based first-line antiretroviral therapy for people with HIV in South Africa: a retrospective cohort study. Lancet HIV. May 2023;10(5):e284–e294. doi:10.1016/S2352-3018(23)00047-4

8. Correa A, Monteiro P, Calixto F, Batista JDL, de Alencar Ximenes RA, Montarroyos UR. Dolutegravir: Virologic response and tolerability of initial antiretroviral regimens for adults living with HIV. PLoS One. 2020;15(8):e0238052. doi:10.1371/journal.pone.0238052

9. Asare K, Sookrajh Y, van der Molen J, et al. Clinical outcomes with second-line dolutegravir in people with virological failure on first-line non-nucleoside reverse transcriptase inhibitor-based regimens in South Africa: a retrospective cohort study. Lancet Glob Health. Feb 2024;12(2):e282–e291. doi:10.1016/S2214-109X(23)00516-8

10. Tschumi N, Lerotholi M, Motaboli L, Mokete M, Labhardt ND, Brown JA. Two-Year Outcomes of Treatment-Experienced Adults After Programmatic Transitioning to Dolutegravir: Longitudinal Data From the VICONEL Human Immunodeficiency Virus Cohort in Lesotho. Clin Infect Dis. Nov 11 2023;77(9):1318–1321. doi:10.1093/cid/ciad390

11. Chu C, Tao K, Kouamou V, et al. Prevalence of Emergent Dolutegravir Resistance Mutations in People Living with HIV: A Rapid Scoping Review. Viruses. Mar 4 2024;16(3)doi:10.3390/v16030399

12. Bhatt N, Ismail N, Magule C, Hussein C, Meque I et al. HIV Drug Resistance Profile in Clients Experiencing Treatment Failure after the Transition To a Dolutegravir-Based First-Line Antiretroviral Treatment Regimen in Mozambique [Abstract 51]. In Proceedings of the InternationalWorkshop on HIV Drug Resistance and Treatment Strategies, Cape Town, South Africa, 20–22 September 2023.

13. Kalata N, Pals S., Bighignoli B, Kabaghe A, Mkungudza J, et al. HIV Drug Resistance to Dolutegravir Is Uncommon among Adults Investigated for Treatment Failure in Malawi [Abstract 53]. In Proceedings of the International Workshop on HIV Drug Resistance and Treatment Strategies, Cape Town, South Africa, 20–22 September 2023.

14. Namayanja G., Watera C, Pais S, Asio J, Ssemwanga D, et al. Cyclic Acquired HIV Drug Resistance to Dolutegravir among People Living with HIV in Uganda. National Remnant Samples Surveillance 2022 [Abstract 50]. In Proceedings of the International Workshop on HIV Drug Resistance and Treatment Strategies, Cape Town, South Africa, 20–22 September 2023.

15. Parikh A, Ochleng C, Almas S, Habib P, Towett J et al, HIV Drug Resistance in the Dolutegravir Era: Preliminary Results from East Africa [Abstract 56]. In Proceedings of the International Workshop on HIV Drug Resistance and Treatment Strategies, Cape Town, South Africa, 20–22 September 2023.

16. Allahna E, Nicole D, Neha S, et al. Brief Report: Virologic Impact of the Dolutegravir Transition: Prospective Results From the Multinational African Cohort Study. J Acquir Immune Defic Syndr. Nov 1 2022;91(3):285–289. doi:10.1097/QAI.0000000000003065

17. HIV Drug Resistance. Brief Report 2024. Geneva: World Health Organization. 2024. Accessed on April 12, 2024 at: https://iris.who.int/bitstream/handle/10665/376039/9789240086319-eng.pdf?sequence=1.

18. Panel on Antiretroviral Guidelines for Adults and Adolescents. Guidelines for the Use of Antiretroviral Agents in Adults and Adolescents with HIV. Department of Health and Human Services. Accessed on November 5, 2023 at: https://clinicalinfo.hiv.gov/en/guidelines/hiv-clinical-guidelines-adult-and-adolescent-arv/whats-new.

19. European AIDS Clinical Society (EACS) Guidelines Version 12.0, October 2023. Accessed on April 12, 2024 at: https://www.eacsociety.org/media/guidelines-12.0.pdf.

20. Cahn P, Madero JS, Arribas JR, et al. Dolutegravir plus lamivudine versus dolutegravir plus tenofovir disoproxil fumarate and emtricitabine in antiretroviral-naive adults with HIV-1 infection (GEMINI-1 and GEMINI-2): week 48 results from two multicentre, double-blind, randomised, non-inferiority, phase 3 trials. Lancet. Jan 12 2019;393(10167):143–155. doi:10.1016/S0140-6736(18)32462-0

21. Cahn P, Sierra Madero J, Arribas JR, et al. Three-year durable efficacy of dolutegravir plus lamivudine in antiretroviral therapy - naive adults with HIV-1 infection. AIDS. Jan 1 2022;36(1):39–48. doi:10.1097/QAD.0000000000003070

22. Llibre JM, Brites C, Cheng CY, et al. Efficacy and Safety of Switching to the 2-Drug Regimen Dolutegravir/Lamivudine Versus Continuing a 3- or 4-Drug Regimen for Maintaining Virologic Suppression in Adults Living With Human Immunodeficiency Virus 1 (HIV-1): Week 48 Results From the Phase 3, Noninferiority SALSA Randomized Trial. Clin Infect Dis. Feb 18 2023;76(4):720–729. doi:10.1093/cid/ciac130

23. Osiyemi O, De Wit S, Ajana F, et al. Efficacy and Safety of Switching to Dolutegravir/Lamivudine Versus Continuing a Tenofovir Alafenamide-Based 3- or 4-Drug Regimen for Maintenance of Virologic Suppression in Adults Living With Human Immunodeficiency Virus Type 1: Results Through Week 144 From the Phase 3, Noninferiority TANGO Randomized Trial. Clin Infect Dis. Sep 29 2022;75(6):975–986. doi:10.1093/cid/ciac036

24. Veras N, Pinho R, Ferreira A, Ferreira AC, Kamiensky B, et al. Monitoring HIV dual-therapy implementation as regimen simplification policy in Brazil. Poster 1058.Conference on Retroviruses and Opportunistic Infections (CROI), February 2023, Seattle, WA, USA.

25. UNAIDS Country Fact Sheets. Haiti. 2021. Accessed May 1, 2023 at: https://www.unaids.org/en/regionscountries/countries/haiti.

26. United Nations Development Program, Human Development Reports, 2022. Accessed March 15, 2023 at: https://hdr.undp.org/data-center/country-insights#/ranks.

27. United Nations Meetings Coverage and Press Releases. Security Council Authorizes Multinational Security Support Mission for Haiti. October 2, 2023. https://press.un.org/en/2023/sc15432.doc.htm.

28. Consolidated guidelines on HIV prevention, testing, treatment, service delivery and monitoring: recommendations for a public health approach. World Health Organization. July 2021. Accessed May 1, 2023 at: https://www.who.int/publications/i/item/9789240031593.

29. Hennessey KA, Leger TD, Rivera VR, et al. Retention in Care among Patients with Early HIV Disease in Haiti. J Int Assoc Provid AIDS Care. Nov/Dec 2017;16(6):523–526. doi:10.1177/2325957417742670

30. Guiteau Moise C, Rivera VR, Hennessey KA, et al. A Successful Model of Expedited Antiretroviral Therapy for Clinically Stable Patients Living With HIV in Haiti. J Acquir Immune Defic Syndr. Sep 1 2018;79(1):70–76. doi:10.1097/QAI.0000000000001725

31. McCluskey SM, Pepperrell T, Hill A, Venter WDF, Gupta RK, Siedner MJ. Adherence, resistance, and viral suppression on dolutegravir in sub-Saharan Africa: implications for the TLD era. AIDS. Dec 15 2021;35(Suppl 2):S127–S135. doi:10.1097/QAD.0000000000003082

32. Pepperrell T, Venter WDF, McCann K, et al. Participants on Dolutegravir Resuppress Human Immunodeficiency Virus RNA After Virologic Failure: Updated Data from the ADVANCE Trial. Clin Infect Dis. Aug 16 2021;73(4):e1008–e1010. doi:10.1093/cid/ciab086

33. Keene CM, Griesel R, Zhao Y, et al. Virologic efficacy of tenofovir, lamivudine and dolutegravir as second-line antiretroviral therapy in adults failing a tenofovir-based first-line regimen. AIDS. Jul 15 2021;35(9):1423–1432. doi:10.1097/QAD.0000000000002936

34. Loosli T, Hossmann S, Ingle SM, et al. HIV-1 drug resistance in people on dolutegravir-based antiretroviral therapy: a collaborative cohort analysis. Lancet HIV. Nov 2023;10(11):e733–e741. doi:10.1016/S2352-3018(23)00228-X

35. Francois K, Van Onacker JD, Jordan MR, et al. First case report of a perinatally HIV-infected infant with HIV resistance to dolutegravir associated with tenofovir/lamivudine/dolutegravir use in mothers. AIDS. Nov 1 2023;37(13):2097–2099. doi:10.1097/QAD.0000000000003653

36. Brown JA, Nsakala BL, Mokhele K, et al. Viral suppression after transition from nonnucleoside reverse transcriptase inhibitor- to dolutegravir-based antiretroviral therapy: A prospective cohort study in Lesotho (DO-REAL study). HIV Med. Mar 2022;23(3):287–293. doi:10.1111/hiv.13189

37. Gupta SK, Post FA, Arribas JR, et al. Renal safety of tenofovir alafenamide vs. tenofovir disoproxil fumarate: a pooled analysis of 26 clinical trials. AIDS. Jul 15 2019;33(9):1455–1465. doi:10.1097/QAD.0000000000002223

38. Mtisi TJ, Ndhlovu CE, Maponga CC, Morse GD. Tenofovir-associated kidney disease in Africans: a systematic review. AIDS Res Ther. Jun 6 2019;16(1):12. doi:10.1186/s12981-019-0227-1

39. Beard N, Hill A. Combined “Test and Treat” Campaigns for Human Immunodeficiency Virus, Hepatitis B, and Hepatitis C: A Systematic Review to Provide Evidence to Support World Health Organization Treatment Guidelines. Open Forum Infect Dis. Feb 2024;11(2):ofad666. doi:10.1093/ofid/ofad666

40. Reaching the 95-95-95 targets: The importance of multi-stakeholder collaboration. International AIDS Society. Accessed at https://www.iasociety.org/sites/default/files/CPP/IAS-CPP-Key-considerations.pdf.

41. Ferreyra C, Moreto-Planas L, Wagbo Temessadouno F, et al. Evaluation of a community-based HIV test and start program in a conflict affected rural area of Yambio County, South Sudan. PLoS One. 2021;16(7):e0254331. doi:10.1371/journal.pone.0254331

42. Ferreyra C, O’Brien D, Alonso B, Al-Zomour A, Ford N. Provision and continuation of antiretroviral therapy during acute conflict: the experience of MSF in Central African Republic and Yemen. Confl Health. 2018;12:30. doi:10.1186/s13031-018-0161-1

